# Comparison of patients diagnosed with intimate partner violence with and without brain injury: Retrospective cross-sectional study in a single, urban, academic health system, 2016-2025

**DOI:** 10.64898/2026.10.21.26366030

**Authors:** Chuan-Ching Tsai, Dorothy A. Kozlowski, Sonya V. Crabtree-Nelson, David Sedillo, Shelby Hoffman Binder, Emoonah McClerklin, Monika Pitzele, Ruchi Fitzgerald, Raj C. Shah

**Affiliations:** Rush Alzheimer’s Disease Center, Rush University Medical Center, Chicago, Illinois, USA; Department of Neuroscience, DePaul University, Chicago, Illinois, USA; Social Work Department, DePaul University, Chicago, Illinois, USA; Policy, Advocacy, and Research Team, The Network: Advocating Against Domestic Violence, Chicago, Illinois, USA; Department of Emergency Medicine, Rush University Medical Center, Chicago, Illinois, USA; PCC Community Wellness Center, Oak Park, Illinois, USA; Department of Family and Preventive Medicine, Rush University Medical Center, Chicago, Illinois, USA.

**Keywords:** brain injury, interpersonal violence, domestic violence, prevalence, co-morbidities, health utilization, emergency room

## Abstract

**Background:** Brain injury (BI) in people who have experienced intimate partner violence (IPV) is underdiagnosed in health care settings. Our project aims to determine how the prevalence and characteristics of people with documented IPV+BI differ from people with IPV alone.

**Methods:** A retrospective cross-sectional study was conducted using electronic health record data at Rush University Medical Center (2016–2025). Adults aged 18 years or older were classified into two groups: IPV with brain injury (IPV+BI) and IPV without brain injury (IPV). Demographics, injury patterns, comorbidities, and pre-diagnosis healthcare utilization between cohorts were compared.

**Results:** In 1,329,460 adult patients from 2016 to 2025 seen at Rush, 2,816 were diagnosed with IPV (prevalence = 0.21%). Also, 486 were concomitantly diagnosed with IPV+BI (17.2% of the IPV population). Most (71%) of initial diagnosis documentation occurred in an Emergency Room encounter. Alcohol use disorder, anemia, chronic liver disease, and vascular diseases were more common in the IPV+BI cohort, compared to IPV alone. Prior to diagnosis, the IPV+BI cohort utilized significantly more acute care resources but less ambulatory usage.

**Discussion:** Electronic health record documentation of IPV with and without BI mainly occurred in the Emergency Room and was lower than anticipated. People with IPV+BI were more likely to have certain co-morbidities and were less likely to utilize health resources before diagnosis. The findings, if confirmed utilizing data from multiple health systems in the Chicagoland region, can support work to advance diagnosis and management of IPV+BI.

**KEYPOINTS:**

- In 1,329,460 adult patients from 2016 to 2025 seen at Rush, 2,816 were diagnosed with IPV (prevalence = 0.21%) which is lower than reported from public health datasets.
- Also, 486 were concomitantly diagnosed with IPV+BI (17.2% of the IPV population).
- The IPV and IPV+BI were initially documented during an emergency room encounter (71% overall, 68% for IPV alone and 93% for IPV+BI) with injury type mainly being unspecified or other head injury rather (74%) rather than traumatic brain injury or strangulation.
- In the IPV+BI cohort, alcohol use disorder, anemia, chronic liver disease, and vascular diseases were significantly more common than the IPV alone.
- Before diagnosis, the IPV+BI cohort utilized more acute care but lacked ambulatory usage.

## 1. Introduction

Intimate partner violence (IPV) in the United States is a significant public health issue with the last nationally representative survey a decade ago concluding that 47.3% of adult women and 44.2% of adult men experienced IPV at some point in their lives [1]. IPV is defined by the United States Centers for Disease Control and Prevention (CDC) as “physical violence, sexual violence, stalking and psychological aggression (including coercive tactics) by a current or former intimate partner” [2].

Brain injury (BI) is a common but underrecognized chronic sequela of IPV. BI can occur due to either traumatic brain injury (TBI) or hypoxic-ischemic brain injury due to strangulation [3]. TBI is defined by the CDC as “a disruption in the normal function of the brain caused by a bump, blow, jolt to the head, or a penetrating head injury” [4]. Closed head TBI is caused by rapid acceleration/deceleration of the head which in turn results in mechanical injury to the brain tissue and vasculature [5]. Secondary injury occurs later due to cellular, chemical, tissue, and blood vessel changes that result in cell death and destruction of brain tissue. In addition to TBI, people who experience IPV may experience hypoxic/ischemic brain injury due to strangulation (defined as the blocking of air or blood flow to and from the head by compressing the neck or torso) [6].

BI may interfere with everything from initial contact with first responders to follow up visits with health care workers. Impaired decision making and memory disruption from BI can interact with many other factors to challenge one’s ability to leave a violent relationship, hold the person causing harm accountable in the legal system, and remember healthcare or social services appointments. It may also affect emotional regulation that can cause safety risks both in the home or while staying in shelter.

Data analysis of the CDC’s National Intimate Partner and Sexual Violence Survey found 3.4% of women and 0.6% of men report being “knocked out after getting hit, slammed against something, or choked” by an intimate partner and that 4.3 million women and 691,000 men in the US have experienced at least one BI [1]. In a 2022 scoping review of 18 articles assessing the prevalence of brain injuries among people living with IPV and seeking social services, BI estimates varied from 28% to 100% [7]. IPV+BI prevalence from health system records has been limited to date. Using data from the National Trauma Data Bank from 2018-2021, concurrent BI was found to be present in 31.1% of IPV cases [8]. Given the calls for more localized data to enable better service delivery to people living with IPV+BI by health and social organizations [9,10], our project aims to provide epidemiological information utilizing electronic health record curated data from an urban academic health system serving communities in the Chicago region, especially on the West Side of the City of Chicago. We utilize this information to describe characteristics of people with documented IPV+BI and how they differ from people with documented IPV alone.

## 2. Methods

### 2.1. General Design

We conducted a retrospective cross-sectional study of people seeking care from January 1, 2016, to December 31, 2025, at a single, urban academic health system who either had a diagnosis of IPV+BI or a diagnosis of IPV alone. The study was determined to be non-human subjects research by the Rush Institutional Review Board.

### 2.2. Data source

We leveraged the Chicago Area Patient-Centered Outcomes Research Network (CAPriCORN) infrastructure at Rush as the main data source. The specifications of the CAPriCORN Common Data Model at Rush are congruent with the PCORnet® Common Data Model (CDM) version 7.0 [11]. CAPriCORN is a partnership between healthcare and research institutions that provides deidentified, deduplicated medical record-based data through a federated harmonized common data model [12,13]. It has been refreshed quarterly at Rush and encompasses various aspects of healthcare, including demographics, encounters, diagnoses, procedures, medications, labs, and patient-reported data. As of December 2025, CAPriCORN at Rush consisted of over 2 million unique patients and their clinical data starting from 2011.

### 2.3. Participants

Patients were eligible if they had an encounter (engagement with a health provider) between January 1, 2016, and December 31, 2025, were diagnosed with IPV, and were 18 years or older at IPV diagnosis. Starting in 2016 enabled focused use of previously published electronic phenotypes based on ICD-10 codes (rather than ICD-9 codes) to identify people diagnosed with IPV with or without concurrent BI (See Appendix, Supplemental Table A and B). [8,14] The index date (analytic time zero) for the IPV+BI group was the date of first IPV visit with a linked BI in an encounter within +/- 180 days of that visit. The index date for the IPV alone cohort was the first IPV visit.

### 2.4. Variables

#### 2.4.1. Outcome variables

The main outcome variables for this study were the prevalence of IPV and IPV+BI. The denominator for both prevalences was the number of unique adult patients seen at Rush during the study period from 2016 to 2025. For the IPV prevalence, the numerator was the count of patients with a recorded IPV diagnostic code but no BI code within six months. For IPV+BI, the numerator was the count of patients with IPV+BI during this timeframe. The prevalence of IPV+BI was divided by the prevalence of IPV only to obtain the prevalence of BI within those diagnosed with IPV.

#### 2.4.2 Cohort Characterization Variables

Demographic characteristics include sex, race, ethnicity, sexual orientation, gender identity, and preferred language which are standard columns in the PCORnet® CDM v7.0 demographics table. In addition, insurance type was obtained from the encounters table. These were categorical variables that only accepted codes from a predetermined list. Clinical characteristics include injury patterns, comorbidities, and healthcare utilization. Injury patterns were analyzed within the IPV+BI group and included the proportion of patients stratified by type of BI based on ICD-10 codes in three categories: TBI, strangulation, and unspecified or other head injury which included scalp laceration and scalp contusion (see Appendix, Supplemental Table B).

Comorbidities were defined as diagnosis made within a year prior to the index IPV visit, were extracted from the diagnosis table and were grouped into 24 common condition categories based on ICD-10 codes (see Appendix, Supplemental Table C). Pre-diagnosis of IPV healthcare utilization required extraction of all encounters within one year prior to the index date from the encounter table. Encounter types were grouped into “ambulatory” (including ambulatory, other ambulatory, or telehealth visits) or “acute care” (including emergency department visits, emergency department with later inpatient admission, inpatient admission, and observation visits).

### 2.5. Analyses

First, we calculated the prevalence outcomes, which were reported in both percentages and the numerator/denominator values. Then, we reported the key demographic characteristics of the overall IPV cohort, the IPV+BI cohort, and the IPV only cohort in terms of both absolute count and percentages. Comparing the IPV+BI versus IPV cohorts, chi-square test (or Fisher’s exact test for expected cell counts < 5) was applied to test the statistical difference of categorical variables (sex, race, ethnicity, sexual orientation, preferred language) and Mann-Whitney U tests was used for continuous variables (age). Among individuals in the IPV+BI cohort, we reported the number and percentage of patients in each injury type category relative to the entire IPV+BI cohort. Patients were only counted once per category. Strangulation and TBI were not mutually exclusive. Patients were assigned to “unspecified/other head injury” only if no strangulation or TBI codes were found. For each comorbid condition, we calculated the number and percentage of individuals in the IPV+TBI and IPV only cohorts. Differences in comorbidity prevalence between cohorts were assessed using chi-squared test or Fisher’s exact test, based on cell counts. Odds ratios (ORs) and 95% confidence intervals (CIs) were calculated to quantify the magnitude of association. For each patient, we calculated binary indicators of any ambulatory and any acute care utilization. The utilization pattern was compared between the IPV+BI and the IPV cohort using chi-square tests, with Holm-Bonferroni correction across the two comparisons.

## 3. Results

### 3.1. Prevalence of People Diagnosed with IPV+BI versus IPV only

From January 1, 2016, to December 31, 2025, there were 1,329,460 unique adult patients seen at Rush. Of these, 2,816 unique patients received an IPV diagnosis, resulting in a prevalence of 0.21%. The prevalence of IPV+BI is 0.037% (486/1,329,460) and the prevalence of BI within IPV was 17.3% (486/2816). The Emergency Room was the most common encounter for a diagnosis of IPV overall (72%). Those who were also diagnosed with BI received this diagnosis in the ER at a rate of 91%. Of all adult patients with an emergency room visit, 0.40% (2,022/510,472) received an IPV diagnosis.

### 3.2 Demographic, Health Utilization, and Clinical Characteristics Differences of People Diagnosed with IPV+BI versus IPV only

The demographic breakdown of the IPV+BI, IPV alone, and the overall IPV cohort is detailed in Table 1. Of this overall cohort, the median age was 31 years old at diagnosis (inter-quartile range: 25-43); 90% were female, 45.2% were African American, and 75.3% were non-Hispanic. The percentage identifying as LGBTQ+ was 3.4% (n=95), and 5.4% (n=153) had a preferred language other than English. For reference, the adults seeking any care at Rush between 2016 and 2025 had a median age of 46 (IQR: 30-66); 55.1% were female; 22.1% were African American; and 74.7% are non-Hispanic. In addition, the overall cohort of adults seen in the Emergency Room was 35% African American. The IPV+BI cohort differed from the IPV alone cohort for the age and race variables. The IPV+BI group was more likely than the IPV alone group to be in the 35 to 54 years range rather than the 18 to 34 years range. Also, IPV+BI was more likely to be documented as African Americans rather than White.

**Table 1.** Demographic Characteristics of Patients with IPV+BI, IPV alone, and the Overall IPV Cohort Absolute counts and percentages within the cohort were reported. Characteristics of all adult patients with any visit and with an emergency room visit are also included.

| Variable | Variable values | Adult Patient Cohort |  |  |  |  |  |
| --- | --- | --- | --- | --- | --- | --- | --- |
|  |  | IPV+BI<br>N=486 | IPV alone<br>N=2,330 | Overall IPV<br>N=2,816 | All<br>N=1,329,460 | All with<br>Emergency Visit<br>N=501,472 | p-value* |
| Age | Median (IQR) | 34 (26-45) | 31 (24-43) | 31 (25-43) | 45 (30-61) | 42 (29-60) | 0.003 |
|  | 18-34 | 252 (51.9%) | 1,370 (58.8%) | 1,622 (57.6%) | 449,524 (33.8%) | 186,387 (37.2%) |  |
|  | 35-44 | 111 (22.8%) | 422 (18.1%) | 533 (18.9%) | 211,982 (15.9%) | 83,521 (16.7%) |  |
|  | 45-54 | 61 (12.6%) | 225 (9.7%) | 286 (10.2%) | 194,971 (14.7%) | 71,277 (14.2%) |  |
|  | 55-64 | 31 (6.4%) | 159 (6.8%) | 190 (6.7%) | 199,993 (15.0%) | 64,957 (13.0%) |  |
|  | 65+ | 31 (6.4%) | 154 (6.6%) | 185 (6.6%) | 272,990 (20.5%) | 95,330 (19.0%) |  |
| Sex† | Female | 440 (90.5%) | 2,102 (90.2%) | 2,542 (90.3%) | 752,293 (56.6%) | 276,111 (55.1%) | 0.828 |
|  | Male | 46 (9.5%) | 228 (9.8%) | 274 (9.7%) | 576,831 (43.4%) | 225,342 (44.9%) |  |
| Race | Asian | <11 | 47 (2.0%) | 54 (1.9%) | 58,391 (4.4%) | 13,536 (2.7%) | <0.001 |
|  | African American | 259 (53.3%) | 1,014 (43.5%) | 1,273 (45.2%) | 281,111 (21.1%) | 169,519 (33.8%) |  |
|  | White | 115 (23.7%) | 751 (32.2%) | 866 (30.8%) | 663,822 (49.9%) | 192,746 (38.4%) |  |
|  | Other | 100 (20.6%) | 483 (20.7%) | 583 (20.7%) | 246,601 (18.5%) | 112,654 (22.5%) |  |
|  | Unknown | <11 | 35 (1.5%) | 40 (1.4%) | 79,535 (6.0%) | 13,017 (2.6%) |  |
| Ethnicity†† | Non-Hispanic | 367 (75.5%) | 1,753 (75.2%) | 2,120 (75.3%) | 1,005,849 (75.7%) | 371,159 (74.0%) | 0.986 |
|  | Hispanic | 114 (23.5%) | 554 (23.8%) | 668 (23.7%) | 263,805 (19.8%) | 120,518 (24.0%) |  |
|  | Other or unknown | <11 | 23 (1.0%) | 28 (1.0%) | 59,806 (4.5%) | 9,795 (2.0%) |  |
| Sexual orientation†† | Heterosexual, other, or unknown | 474 (97.5%) | 2,247 (96.4%) | 2,721 (96.6%) | 1,314,021 (98.8%) | 495,273 (98.8%) | 0.456 |
|  | LGBTQ+ | 12 (2.5%) | 83 (3.6%) | 95 (3.4%) | 15,439 (1.2%) | 6,199 (1.2%) |  |
| Preferred language†† | English | 451 (92.8%) | 2,167 (93.0%) | 2,618 (93.0%) | 1,127,914 (84.8%) | 442,357 (88.2%) | 0.105 |
|  | Spanish | 17 (3.5%) | 86 (3.7%) | 103 (3.7%) | 78,313 (5.9%) | 35,447 (7.1%) |  |
|  | Other or unknown | 18 (3.7%) | 77 (3.3%) | 95 (3.4%) | 123,233 (9.3%) | 23,668 (4.7%) |  |
| Insurance | Medicaid | 152 (31.3%) | 798 (34.2%) | 950 (33.7%) | 143,914 (10.8%) | 82,852 (16.5%) | 0.158 |
|  | Medicare | 49 (10.1%) | 233 (10.0%) | 282 (10.0%) | 229,856 (17.3%) | 85,375 (17.0%) |  |
|  | Private | 225 (46.3%) | 1,085 (46.6%) | 1,310 (46.5%) | 771,218 (58.0%) | 243,036 (48.5%) |  |
|  | Other or unknown | 60 (12.3%) | 214 (9.2%) | 274 (9.7%) | 184,472 (13.9%) | 90,209 (18.0%) |  |

\*p-value was calculated comparing the IPV+BI to the IPV alone group. † 336 patients in the "All" cohort and 19 in the "All with Emergency Visit" cohort with unknown sex. †† Categories with low cell counts were combined for privacy reasons; however, statistical tests were run based on original categorization.

Among the 486 patients with IPV+BI, 14% were diagnosed with TBI, 12% with strangulation, and the majority (74%) were diagnosed with unspecified or other head injury. Common comorbidities in the IPV+BI and IPV alone cohorts are shown in Table 2. IPV+BI patients had a statistically higher prevalence of alcohol use disorder (5.8% vs 2.6%), chronic liver disease/cirrhosis (4.7% vs 2.4%), nutritional deficiency anemia (6.0% vs 3.0%), cerebrovascular disease/stroke (4.3% vs 1.9%), and peripheral vascular disease (3.3% vs 1.8) (all p < 0.05). Also, there was a higher tendency for IPV+BI cohorts to exhibit more suicidal/homicidal ideation or attempt (p=0.054). Other comorbidities above 10% but similar between the two cohorts were hypertension, anxiety, asthma, depression, and obesity.

**Table 2.** Prevalence of Comorbidities by Cohort (IPV+BI vs IPV alone). Comorbidities identified within 365-day pre-index period. Values are n (%), sorted by prevalence in the IPV+BI cohort. ORs (95% CIs) compare IPV+BI vs IPV alone. Significant results (p<0.05) denoted with an asterisk (*).

| Category | IPV+BI<br>(N=486) | IPV alone<br>(N=2,330) | OR (95% CI) | p-value |
| --- | --- | --- | --- | --- |
| Hypertension | 74 (15.2%) | 291 (12.5%) | 1.26 (0.95-1.66) | 0.10 |
| Anxiety | 71 (14.6%) | 311 (13.3%) | 1.11 (0.84-1.47) | 0.46 |
| Asthma | 58 (11.9%) | 224 (9.6%) | 1.27 (0.94-1.73) | 0.12 |
| Depression | 56 (11.5%) | 252 (10.8%) | 1.07 (0.79-1.46) | 0.65 |
| Obesity | 45 (9.3%) | 248 (10.6%) | 0.86 (0.61-1.20) | 0.36 |
| Chronic pain | 44 (9.1%) | 190 (8.2%) | 1.12 (0.80-1.58) | 0.51 |
| Hyperlipidemia | 37 (7.6%) | 158 (6.8%) | 1.13 (0.78-1.64) | 0.51 |
| Substance use disorder | 32 (6.6%) | 120 (5.2%) | 1.30 (0.87-1.94) | 0.20 |
| Nutritional deficiency anemia* | 29 (6.0%) | 70 (3.0%) | 2.05 (1.31-3.20) | 0.001 |
| Alcohol use disorder * | 28 (5.8%) | 60 (2.6%) | 2.31 (1.46-3.66) | <0.001 |
| Suicidal/homicidal ideation or attempt | 24 (4.9%) | 74 (3.2%) | 1.58 (0.99-2.54) | 0.054 |
| Diabetes without complication | 23 (4.7%) | 119 (5.1%) | 0.92 (0.58-1.46) | 0.73 |
| Chronic liver disease and cirrhosis * | 23 (4.7%) | 55 (2.4%) | 2.05 (1.25-3.38) | 0.004 |
| Cerebrovascular disease and stroke * | 21 (4.3%) | 44 (1.9%) | 2.35 (1.38-3.98) | 0.001 |
| Coronary ischemic heart disease | 19 (3.9%) | 86 (3.7%) | 1.06 (0.64-1.76) | 0.82 |
| Post Traumatic Stress Disorder | 18 (3.7%) | 67 (2.9%) | 1.30 (0.76-2.21) | 0.33 |
| Peripheral vascular disease* | 16 (3.3%) | 43 (1.8%) | 1.81 (1.01-3.24) | 0.043 |
| Diabetes with complication | 16 (3.3%) | 78 (3.3%) | 0.98 (0.57-1.70) | 0.95 |
| Chronic Obstructive Pulmonary Disease | 15 (3.1%) | 46 (2.0%) | 1.58 (0.88-2.86) | 0.13 |
| Congestive heart failure | 13 (2.7%) | 57 (2.4%) | 1.10 (0.60-2.02) | 0.77 |
| Schizophrenia and psychotic disorders | 11 (2.3%) | 59 (2.5%) | 0.89 (0.46-1.71) | 0.73 |
| Chronic kidney disease | 11 (2.3%) | 54 (2.3%) | 0.98 (0.51-1.88) | 0.94 |
| Any cancer (solid tumor or hematologic) | 6 (1.2%) | 31 (1.3%) | 0.93 (0.38-2.23) | 0.87 |
| HIV/AIDS | 3 (0.6%) | 11 (0.5%) | 1.31 (0.36-4.71) | 0.72 |

Healthcare utilization differed between cohorts in the year prior to diagnosis (Figure 1). Patients with IPV+BI had lower ambulatory utilization than IPV patients, with fewer patients having any ambulatory encounters (31.9% vs 41.6%, Holm adjusted-p <0.001). In contrast, acute care utilization was higher in the IPV+BI cohort, with a greater percentage with any acute care encounter (50.8% vs 36.2%, Holm adjusted-p <0.001).

**Figure 1.**
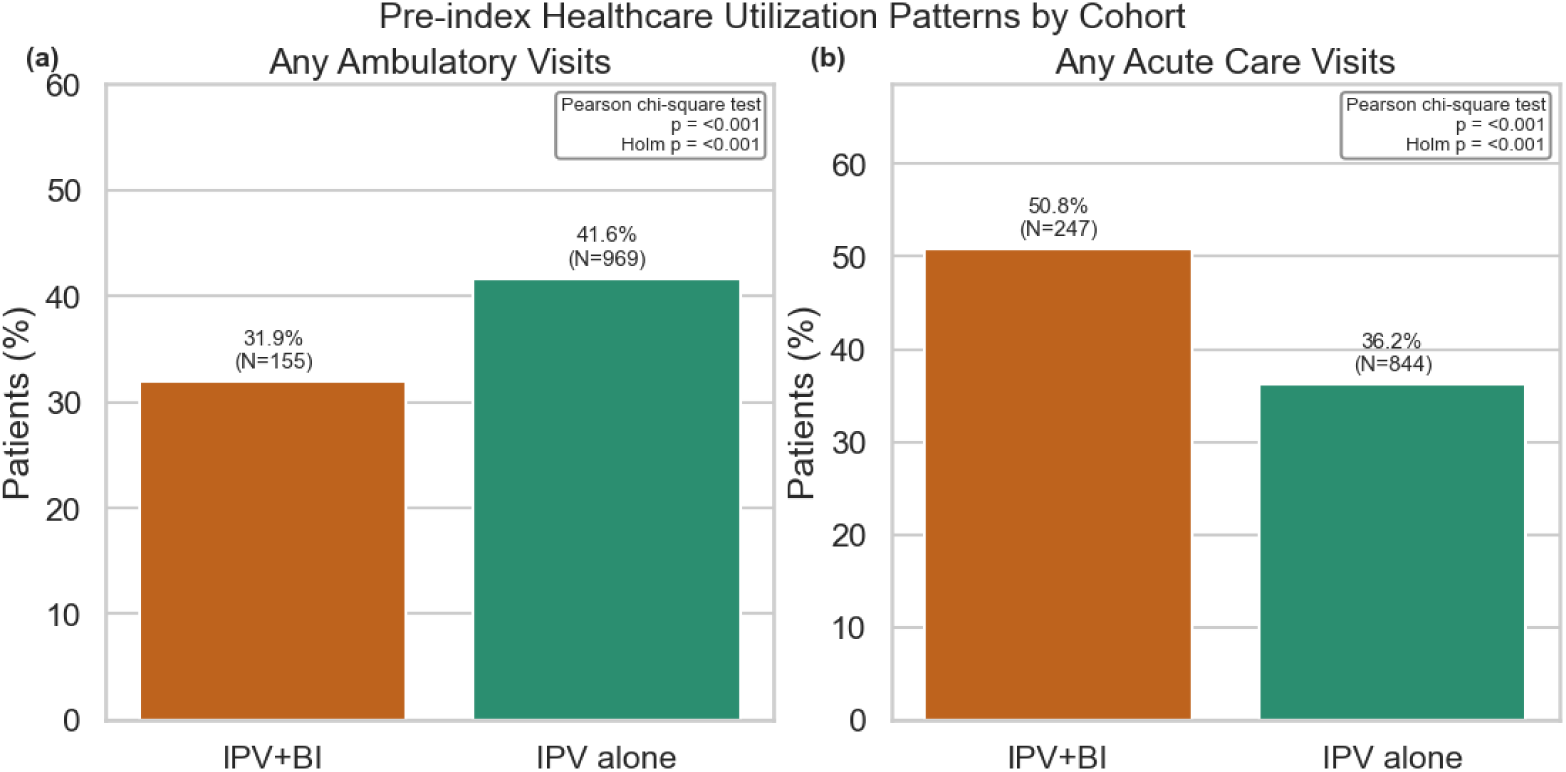
Healthcare utilization pattern by cohort (IPV+BI vs IPV). Panels (a) and (b) show the proportion and absolute count of participants with any ambulatory and acute care visits, respectively; Annotations show cohort-specific summary statistics. Between-cohort differences were tested using chi-square tests, with Holm-Bonferroni correction across the two comparisons.

## 4. Discussion

Using a curated medical record-based data model at a single, urban, academic health system, we characterized the prevalence of IPV+BI versus IPV alone and then identified between group differences. IPV prevalence in patients seen at Rush from 2016 to 2025 was 0.21% and only 0.037% were concomitantly diagnosed with IPV+BI. The prevalence of BI in the IPV group was 17.3%. In both cohorts, most diagnoses were made during emergency room encounters (0.40% with IPV+BI in all adults with at least one emergency room visit). Compared to the IPV alone cohort, the IPV+BI cohort was (1) older in age and more likely to be African American, (2) utilized more acute care but lacked ambulatory care visits prior to initial IPV diagnosis, and (3) had greater presence of alcohol use disorder, nutritional deficiencies anemia, chronic liver disease, and vascular diseases.

In this electronic medical record cohort, IPV was less prevalent as compared to non-health system-based surveys [1]. The National Hospital Ambulatory Medical Care Survey (between 2002-2015) reported an IPV prevalence of 0.34% of all emergency department visits (for patients 15 years old and above) while the National Electronic Injury Surveillance System – All Injury Program (between 2005-2013) reported an IPV prevalence of 0.6%; both of which are higher than that reported here [15–17]. This difference with other health system-based estimates could be due to the current study using a denominator of all unique adult health system visits rather than just focusing on emergency room encounters. The estimate of IPV when the denominator is all patients with an emergency room visit is 0.40% which is in line with these and other recent studies [18]. Improving documentation in other ambulatory encounters may be of benefit [19]; however, yields may be lower based on underlying pre-screening probabilities as compared to the emergency room. In those diagnosed with IPV, the incidence of brain injury was 17.3%, comparable to 12.5% from another individual health care system in the area [20]. However, this estimate was significantly lower than 80% measured in studies where brain injury incidence is examined in social service or community settings [21,22].

Potential explanations for under reporting may include clinicians not putting codes in for IPV and the hesitancy of some patients living with IPV to disclose IPV issues with the care team [14,23]. Reasons include clinicians’ time constraint [14], inadequate documentation reimbursement [14], the unwillingness of patients to report IPV [24], and patient’s fear of abusive partner’s retaliation [25]. In addition, electronic health record workflows preventing seamless and transparent viewing of physician and nurse/social worker documentation could limit transmission of information from IPV screening done by certain team members to physicians involved in diagnostic, treatment, and coding documentation.

This study also demonstrated that when BI diagnoses were made in a health care center, the BI diagnosis of “unspecified” or “other head injury” was documented in approximately 75% rather than more specific codes associated with TBI or strangulation. Physicians may be hesitant to use a more specific code for TBI if there is no visible scalp injury and a normal computed tomography scan of the brain is present. Strangulation is not often disclosed unless specifically asked since people experiencing IPV with BI may not realize that strangulation could be associated with brain injury and symptomatology. In addition, child custody/welfare involvement could generate hesitancy in documenting BI in a medical record. Screening tools for identifying brain injury from both head trauma and strangulation exist for IPV [26] and could facilitate more specific documentation of BI.

The demographics of the IPV+BI cohort were comparable to IPV alone, except for race and age. Even though the race findings may be influenced by local context in patients seeking care in the non-trauma center emergency room of a single institution, some [27] but not all [8] research has pointed to African Americans experiencing IPV being at elevated risk for BI. The median age for reporting IPV+BI seems older than what would be anticipated if the analysis had not been restricted to people over the age of 18 as there may be a bimodal distribution.

In our study, the IPV+BI group utilized acute care significantly more than the IPV group in the year prior to diagnosis. Interestingly, their usage of ambulatory care prior to diagnosis in the IPV+BI group is significantly less than the IPV alone group. This could be due to the people inflicting harm often interfering with the ability of people living with IPV+BI to obtain medical care [28]. Since a lack of ambulatory care could lead to poorer overall health, mechanisms to support ongoing health care engagement are needed. Forensic programs in emergency departments where training of nurses in trauma-informed care and screening for IPV has been shown to improve care of persons experiencing sexual assault [29–32]. Expansion of the programs to enable better connections to primary care and social services may be an opportunity for advancement of trauma informed care [33], especially if supported by 24-hour social work coverage in the emergency room focused on IPV [34].

IPV alone is associated with increased risk of a variety of physical and mental health conditions both as a direct consequence of violence and in the long-term [35]. In the current study, comorbidities that were common to both cohorts included hypertension, anxiety, asthma, depression, and obesity which have been previously documented [35,36]. Those with IPV+BI had significantly higher comorbidities of alcohol use disorder, chronic liver disease/cirrhosis, anemia, cerebrovascular disease/stroke, peripheral vascular disease, and potentially more suicidal/homicidal ideation or attempts. These comorbidities may reflect multiple interacting factors including trauma, psychiatric comorbidity, substance use, and the sequelae of brain injury.

One strength of the paper is the use of CAPriCORN Common Data Model. With structured data and the use of standardized codes, it offers a significant advantage over chart review, primarily in its ability to rapidly process large datasets. Thereby, it is possible to analyze an expanded patient cohort across greater temporal scales. One limitation of our study is that it only represents information from a single health system. Since CAPriCORN is implemented in several institutions in the Greater Chicago area, work that is underway is to distribute our query to all CAPriCORN sites, to expand population coverage temporally and spatially and to refine local estimates of IPV+BI prevalence. Findings from regional work could support deployment of tailored resources and programs to increase access to medical and social support services for people living with IPV+BI. Effective IPV along with BI screening and referral in emergency departments can help survivors leave violent relationships [37] and is recommended for women of reproductive age by the US Preventative Services Taskforce [38].

## Conclusion

While the documentation of IPV and BI is low, people with documented IPV+BI were more likely to be older and African American in this cohort. Pre-documentation access to ambulatory care was less than emergency and inpatient care and comorbid conditions such as alcohol use disorder, anemia, chronic liver disease, and vascular diseases were more common. If these findings are verified in a larger study, they could facilitate improving screening and treatment by practitioners of emergency medicine, social workers, and service providers in addition to facilitating access to medical and social services for people living with IPV+BI.

## Author Contribution

CT, DAK, SVCN, DS, SHB, and RCS all participated in the design of the project and the critical writing and editing of the manuscript. CT and RCS drafted the manuscript. CT conducted data analyses. CT, RCS, SVCN, DS, SHB, EM, MP, RF, and DAK participated in the critical editing of the manuscript and approved the submission of the final manuscript. CT and DAK both contributing to being co-first authors on this manuscript.

## CRediT authorship contribution statement

**Chuan-Ching Tsai:** Data curation; Formal analysis; Methodology; Visualization; Writing – original draft; Writing – review and editing; **Dorothy A. Kozlowski:** Conceptualization; Funding acquisition; Methodology; Project administration; Resources; Supervision; Validation; Writing – review and editing; **Sonya V. Crabtree-Nelson:** Conceptualization; Methodology; Validation; Visualization; Writing – review and editing; **David Sedillo:** Project administration; Resources; Supervision; Writing – review and editing; **Shelby Hoffman Binder:** Conceptualization; Methodology; Validation; Writing – review and editing; **Emoonah McClerklin:** Validation; Writing – review and editing; **Monika Pitzele:** Validation; Writing – review and editing; **Ruchi Fitzgerald:** Validation; Visualization; Writing – review and editing; **Raj C. Shah:** Conceptualization; Funding acquisition; Investigation; Methodology; Project administration; Resources; Supervision; Validation; Writing – original draft; Writing – review and editing;

## Declaration of generative AI and AI-assisted technologies in the writing process

Generative AI was used in writing SQL and Python codes and grammar checks in the writing process. All AI generated outputs were reviewed by the author.

## Acknowledgements

The authors recognize the significant contributions made by people living with IPV and BI that composed the “Survivor Advisory Group” organized by The Network: Advocating Against Domestic Violence, who consulted on the research questions and analysis of the data in our study and shared powerful lived experience with the research team. The authors acknowledge the clinicians, staff, and patients at Rush involved in the healthcare documented in the electronic health records that were curated into the Rush instance of the PCORnet® Common Data Model.

## Appendix A: Supplementary data

Supplementary materials for this article can be found online in a separate document.

## Data availability

The data cannot be shared given it represents curated patient electronic medical record data. Inquiries regarding collaborative work may be submitted to Raj C. Shah, MD at. Analytic code utilized for this work is publicly available at https://github.com/cctsairush/BI-IPV-Rush-only.

## Appendix: Supplemental Tables and Figures

**Supplemental Table A.**
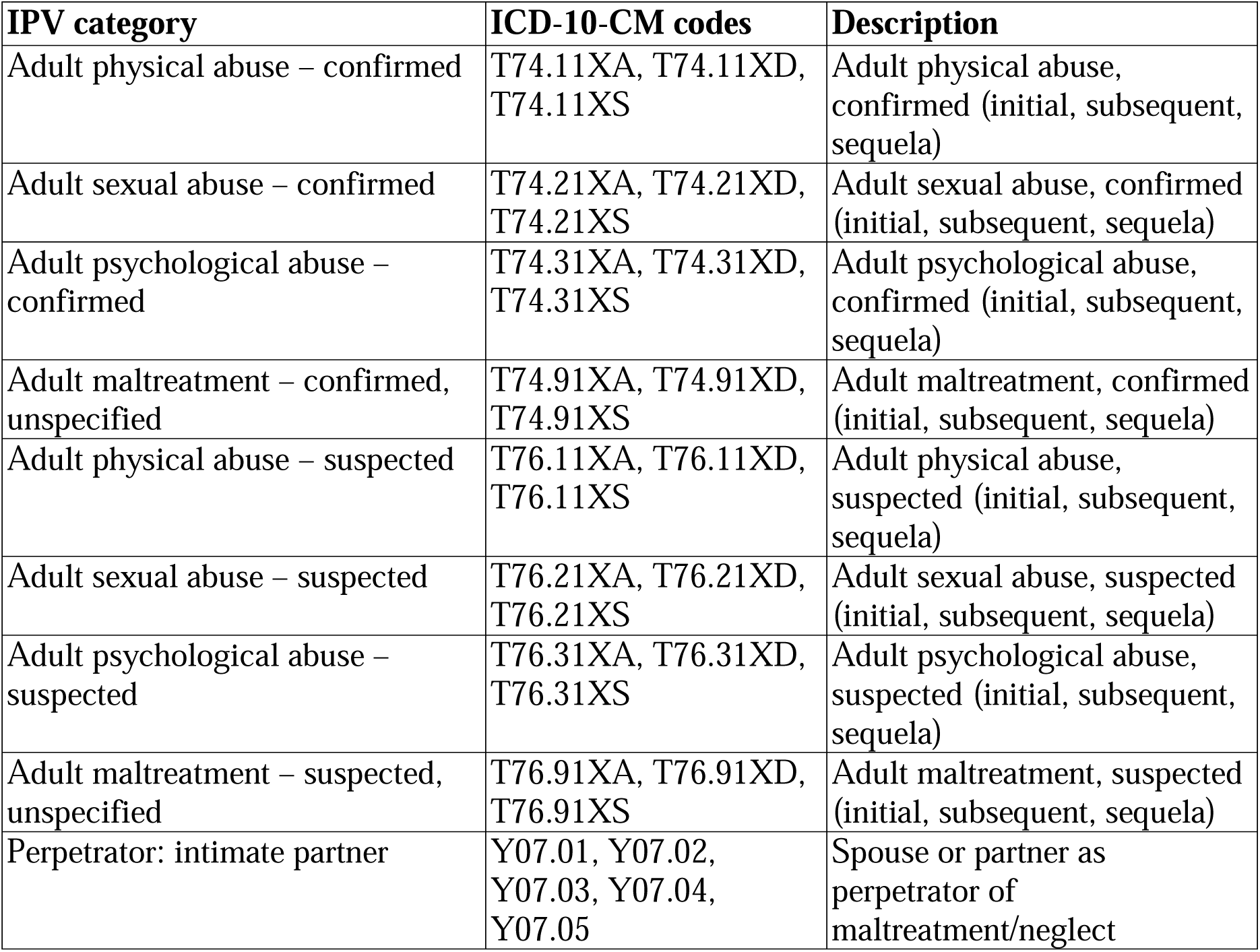
Diagnostic (ICD-10) Codes Used to Identify Intimate Partner Violence (IPV)

| **IPV category** | **ICD-10-CM codes** | **Description** |
| --- | --- | --- |
| Adult physical abuse – confirmed | T74.11XA, T74.11XD, T74.11XS | Adult physical abuse, confirmed (initial, subsequent, sequela) |
| Adult sexual abuse – confirmed | T74.21XA, T74.21XD, T74.21XS | Adult sexual abuse, confirmed (initial, subsequent, sequela) |
| Adult psychological abuse – confirmed | T74.31XA, T74.31XD, T74.31XS | Adult psychological abuse, confirmed (initial, subsequent, sequela) |
| Adult maltreatment – confirmed, unspecified | T74.91XA, T74.91XD, T74.91XS | Adult maltreatment, confirmed (initial, subsequent, sequela) |
| Adult physical abuse – suspected | T76.11XA, T76.11XD, T76.11XS | Adult physical abuse, suspected (initial, subsequent, sequela) |
| Adult sexual abuse – suspected | T76.21XA, T76.21XD, T76.21XS | Adult sexual abuse, suspected (initial, subsequent, sequela) |
| Adult psychological abuse – suspected | T76.31XA, T76.31XD, T76.31XS | Adult psychological abuse, suspected (initial, subsequent, sequela) |
| Adult maltreatment – suspected, unspecified | T76.91XA, T76.91XD, T76.91XS | Adult maltreatment, suspected (initial, subsequent, sequela) |
| Perpetrator: intimate partner | Y07.01, Y07.02, Y07.03, Y07.04, Y07.05 | Spouse or partner as perpetrator of maltreatment/neglect |

**Supplemental Table B.** Diagnostic (ICD-10) Codes for Brain Injury (BI) and Injury Type.

| **ICD-10 CM Codes** | **Brain Injury Category** | **Brain Injury Type** |
| --- | --- | --- |
| F07.81, S06.0 | Concussion & post-concussive syndrome | Traumatic brain injury |
| S06.1 – S06.6 | Skull fracture, Intracranial hemorrhage, intracranial injury, and edema | Traumatic brain injury |
| I60.9 | Subarachnoid hemorrhage | Traumatic brain injury |
| S09.90XA | Unspecified head injury | Unspecified or other head injury |
| S00.03XA | Scalp contusion | Unspecified or other head injury |
| S01.01XA | Scalp laceration | Unspecified or other head injury |
| T71.193A | Strangulation | Strangulation |

**Supplemental Table C.** Diagnostic (ICD-10) Codes Used to Identify Selected Common Chronic Conditions as Comorbidities.

| **Comorbidity category** | **ICD-10-CM codes included** |
| --- | --- |
| HIV/AIDS | B20, Z21, O98.71*, O98.72, O98.73 |
| All cancer (malignant neoplasms) | C16.*, C18.*-C20, C22.*, C25.*, C34.*, C50.*, C53.*, C56.*, C61, C64.*, C65.*, C81.*-C85.*, C90.*, C91.*-C95.* |
| Nutritional deficiency anemia | D50.*-D53.* |
| Diabetes without complication | E08.9, E09.9, E10.9, E11.9, E13.9 |
| Diabetes with complication | E08.*, E09.*, E10.*, E11.*, E13.* excluding *.9 codes (i.e., excluding E08.9/E09.9/E10.9/E11.9/E13.9) |
| Obesity | E66.* |
| Hyperlipidemia | E78.* |
| Alcohol use disorder | F10.* |
| Substance use disorder | F11.*-F19.* |
| Schizophrenia & psychotic disorders | F20.*, F21, F22, F23, F24, F25.*, F28, F29 |
| Depression | F32.*, F33.* |
| Anxiety | F40.*, F41.* |
| Post-Traumatic Stress Disorder | F43.1* |
| Suicidal/homicidal ideation or attempt | R45.850, R45.851, T14.91, Z91.5 |
| Chronic pain | G89.2*, G89.4 |
| Cerebrovascular disease / stroke | G45.*, I60.*-I69.*, I65.*-I68.*, I67.* |
| Hypertension | I10, I11.*, I12.*, I13.*, I15.*, I16.* |
| Congestive heart failure | I09.81, I11.0, I13.0, I13.2, I50.* |
| Coronary ischemic heart disease | I20.*-I25.* |
| Peripheral vascular disease | I70.*-I79.*, plus diabetes-PVD codes E08.51, E09.51, E10.51, E11.51, E13.51 |
| Chronic Obstructive Pulmonary Disease | J41.*-J44.* |
| Asthma | J45.* |
| Chronic liver disease & cirrhosis | K70.*-K77.* |
| Chronic kidney disease | N18.* |

## References

[1] Leemis RW, Friar N, Khatiwada S, et al. The National Intimate Partner and Sexual Violence Survey: 2016/2017 Report on Intimate Partner Violence. Atlanta, GA: National Center for Injury Prevention and Control, Centers for Disease Control and Prevention; 2022.

[2] Breiding M, Basile K, Smith S, et al. Intimate Partner Violence Surveillance: Uniform Definitions and Recommended Data Elements. Atlanta, GA: National Center for Injury Prevention and Control, Centers for Disease Control and Prevention; 2015.

[3] Sheridan DJ, Nash KR. Acute injury patterns of intimate partner violence victims. Trauma Violence Abus. 2007;8:281–9. 10.1177/1524838007303504.

[4] Marr AL. Central Nervous System Injury Surveillance Data Submission Standards—2002. Centers for Disease Control and Prevention National Center for Injury Prevention and Control; 2004.

[5] Giza CC, Hovda DA. The new neurometabolic cascade of concussion. Neurosurgery. 2014;75:S24–S33. 10.1227/NEU.0000000000000505.

[6] Monahan K, Bannon S, Dams-O’Connor K. Nonfatal strangulation (NFS) and intimate partner violence: a brief overview. J Fam Viol. 2022;37:75–86. 10.1007/s10896-020-00208-7.

[7] Campbell JK, Joseph A-LC, Rothman EF, Valera EM. The prevalence of brain injury among survivors and perpetrators of intimate partner violence and the prevalence of violence victimization and perpetration among people with brain injury: a scoping review. Curr Epidemiol Rep. 2022;9:290–315. 10.1007/s40471-022-00302-y.

[8] Ramesh ER, Yue JK, Manley GT, Tarapore PE, DiGiorgio AM. Epidemiology of intimate partner and domestic violence–related traumatic brain injury in the United States, 2018 to 2021: A National Trauma Data Bank Cohort Analysis of 3891 Patients. Neurosurgery. 2024:1–13. 10.1227/neu.0000000000002983.

[9] Basile KC, Smith SG, Friar NW. Defining intimate partner violence. In: Connie Mitchell C, Miller E, McCaw B, Alexander KA, editors. Intimate Partner Violence, second edition, Oxford: Oxford University Press; 2025, p.3–16. 10.1093/oso/9780197758991.003.0001.

[10] United States Government Accountability Office. Domestic Violence: Improved Data Needed to Identify the Prevalence of Brain Injuries among Victims GAO-20-534. Published: Jun 12, 2020. Publicly Released: Jun 12, 2020.

[11] PCORnet®. PCORnet® Common Data Model. https://pcornet.org/data/common-data-model/; 2025 [Accessed 13 October 2026].

[12] Kho AN, Hynes DM, Goel S, et al. CAPriCORN: Chicago area patient-centered outcomes research network. J Am Med Inform Assoc. 2014;21(4):607–11. 10.1136/amiajnl-2014-002827.

[13] Hasjim BJ, Mohammadi M, Balbale SN, et al. High hospitalization rates and risk factors among frail patients with cirrhosis: A 10-year population-based cohort study. Clin Gastroenterol Hepatol. 2025;23(7):1152–1163. 10.1016/j.cgh.2024.08.044.

[14] Rebbe R, Adhia A, Eastman AL, et al. The measurement of intimate partner violence using International Classification of Diseases diagnostic codes: a systematic review. Trauma Violence Abuse. 2023;24(4):2165–80. 10.1177/15248380221090977.

[15] Boyce SC, Silverman JG. Epidemiology of intimate partner violence. In: Connie Mitchell C, Miller E, McCaw B, Alexander KA, editors. Intimate Partner Violence, second edition, Oxford: Oxford University Press; 2025, p.17–56 10.1093/oso/9780197758991.003.0002.

[16] Khurana B, Hines DA, Johnson BA, et al. Injury patterns and associated demographics of intimate partner violence in men presenting to U.S. emergency departments. Aggress Behav. 2022;48(3):298–308. 10.1002/ab.22007.

[17] Khurana B, Bayne HN, Temple J, et al. Comparative analysis of injuries related to self-harm, assault, and intimate partner violence: insights from U.S. emergency departments (2005-2021). Inj Prev. 2026;32(5):758–764. 10.1136/ip-2024-045435.

[18] Akinyemi O, Fasokun M, Thompson T, et al. Socioeconomic determinants and mental health correlates of intimate partner violence-related emergency department visits among women in California. Int J Gynaecol Obstet. 2026. Epub ahead of print. 10.1002/ijgo.71154.

[19] Lenert L, Rheingold AA, Simpson KN, et al. Electronic health record–based screening for intimate partner violence: A Cluster Randomized Clinical Trial. JAMA Netw Open. 2024;7(8):e2425070. 10.1001/jamanetworkopen.2024.25070.

[20] Sachdeva AS, Bell A, Furst DJ, et al. A traumatic brain injury prescreening tool for intimate partner violence patients using initial clinical reports and machine learning. AMIA Jt Summits Transl Sci Proc. 2024;2024:401–8.

[21] Nemeth JM, Decker C, Ramirez R, et al. Partner-inflicted brain injury: intentional, concurrent, and repeated traumatic and hypoxic neurologic insults. Brain Sci. 2025;15(5):524. 10.3390/brainsci15050524.

[22] Esopenko C, Jain D, Hicks AJ, et al. Characterizing intimate partner violence-related head trauma in community-recruited women. J Head Trauma Rehabil. 2025;40(6):e514–23. 10.1097/HTR.0000000000001062.

[23] Olive P. Intimate partner violence and clinical coding: issues with the use of the International Classification of Disease (ICD-10) in England. J Health Serv Res Policy. 2018;23(4):212–21. 10.1177/1355819618781413.

[24] Heron RL, Eisma MC. Barriers and facilitators of disclosing domestic violence to the healthcare service: A systematic review of qualitative research. Health Soc Care Community. 2021;29(3):612–30. 10.1111/hsc.13282.

[25] Randell KA, Ragavan MI, Query LA, et al. Intimate partner violence and the pediatric electronic health record: A Qualitative Study. Acad Pediatr. 2022;22(5):824–832. 10.1016/j.acap.2021.08.013.

[26] Clarke ADA, Copas C, Hannon O, et al. Detecting a hidden pandemic: the current state and future direction of screening and assessment tools for intimate partner violence-related brain injury. Neurosci Biobehav Rev. 2024;167:105912. 10.1016/j.neubiorev.2024.105912.

[27] Banks ME, Ackerman RJ. Head and brain injuries experienced by African American women victims of intimate partner violence. Women & Therapy. 2002;25(3/4):133. 10.1300/J015v25n03_10.

[28] McCloskey LA, Williams CM, Lichter E, et al. Abused women disclose partner interference with health care: an unrecognized form of battering. J Gen Intern Med. 2007;22(8):1067–72. 10.1007/s11606-007-0199-z.

[29] Fehler-Cabral G, Campbell R, Patterson D. Adult sexual assault survivors’ experiences with sexual assault nurse examiners (SANEs). J Interpers Violence. 2011;26(18):3618–39. 10.1177/0886260511403761.

[30] Campbell R, Bybee D, Townsend SM, et al. The impact of sexual assault nurse examiner programs on criminal justice case outcomes: a multisite replication study. Violence Against Women. 2014;20(5):607–625. 10.1177/1077801214536286.

[31] Hollender M, Almirol E, Meyer M, et al. Sexual assault nurse examiners lead to improved uptake of services: a cross-sectional study. West J Emerg Med. 2023;24(5):974–82. 10.5811/westjem.59514.

[32] Campbell R, Patterson D, Lichty LF. The effectiveness of sexual assault nurse examiner (SANE) programs: a review of psychological, medical, legal, and community outcomes. Trauma Violence Abuse. 2005;6(4):313–29. 10.1177/1524838005280328.

[33] Tiyyagura G, Abeyaratne D, Asnes A, et al. Trauma- and violence-informed care practices in the emergency department for survivors of intimate partner violence. JAMA Netw Open. 2026;9(3):e260034. doi:10.1001/jamanetworkopen.2026.0034.

[34] Liao LZ, Bian Y, Brown, OE, Geynisman-Tan J. Barriers and facilitators for intimate partner violence intervention in adult emergency departments in one city. J Fam Viol. 2026;41:1171–81. 10.1007/s10896-024-00776-y.

[35] McCaw B, Mitchell C, Miller E, Alexander KA. Key Considerations regarding health impacts of intimate partner violence. In: Connie Mitchell C, Miller E, McCaw B, Alexander KA, editors. Intimate Partner Violence, second edition, Oxford: Oxford University Press; 2025, p.271–5. 10.1093/oso/9780197758991.003.0010.

[36] Guler A, Coker AL. Intimate partner violence in ambulatory care settings. In: Connie Mitchell C, Miller E, McCaw B, Alexander KA, editors. Intimate Partner Violence, second edition, Oxford: Oxford University Press; 2025, p.276–302. 10.1093/oso/9780197758991.003.0011.

[37] Krasnoff M, Moscati R. Domestic violence screening and referral can be effective. Ann Emerg Med. 2002;40(5):485–92. 10.1067/mem.2002.128872.

[38] US Preventive Services Task Force; Silverstein M, Wong JB, et al. Screening for intimate partner violence and caregiver abuse of older or vulnerable adults: US Preventive Services Task Force recommendation statement. JAMA. 2025;334(4):329–338. 10.1001/jama.2025.9009.

